## Supplementary Information for "The importance of non-pharmaceutical interventions during the COVID-19 vaccine rollout"

### Supplementary Material for *The importance of non-pharmaceutical interventions during the COVID-19 vaccine rollout*

#### Contents

**1 Constant Rate Model** **2**

**2 Symptomatic cases** **3**

**3 Calibration - Posterior Distributions** **5**

**4 Real Vaccination Rollout** **6**

**5 Reopening Scenarios** **7**

**6 Sensitivity Analysis: Behavioural Parameter $\gamma$** **9**

**7 $R_0$ calculation** **10**

### 1 Constant Rate Model

In the main text, we presented results only for the dynamic rate model in which the behavioural transitions are modulated by the fraction of vaccinated population and the number of deaths per $100,000$ in the previous time step. Here, we report in Figure 1 also the behavioural parameters space exploration for simpler constant rate model, in which transitions from and towards the non-compliant compartments are regulated by constant parameters. We explore over a grid of $(\gamma, \alpha)$ pairs, the fraction of averted deaths with respect to a baseline simulation without vaccine (and thus no behaviour change triggered by the vaccination). For comparison, we display in figure also the results presented in the main text for the dynamic rate model. As expected, the overall behaviour of the model is confirmed. For a fixed $\gamma$, the fraction of averted deaths reduces for increasing values of $\alpha$, hinting that a stronger behavioural response causes an additional waste of the benefit brought by the vaccine. Conversely, for a fixed $\alpha$, the fraction of averted deaths increases for increasing values of $\gamma$. Indeed, in these cases non-compliant individuals turn back faster to COVID-safe behaviour.

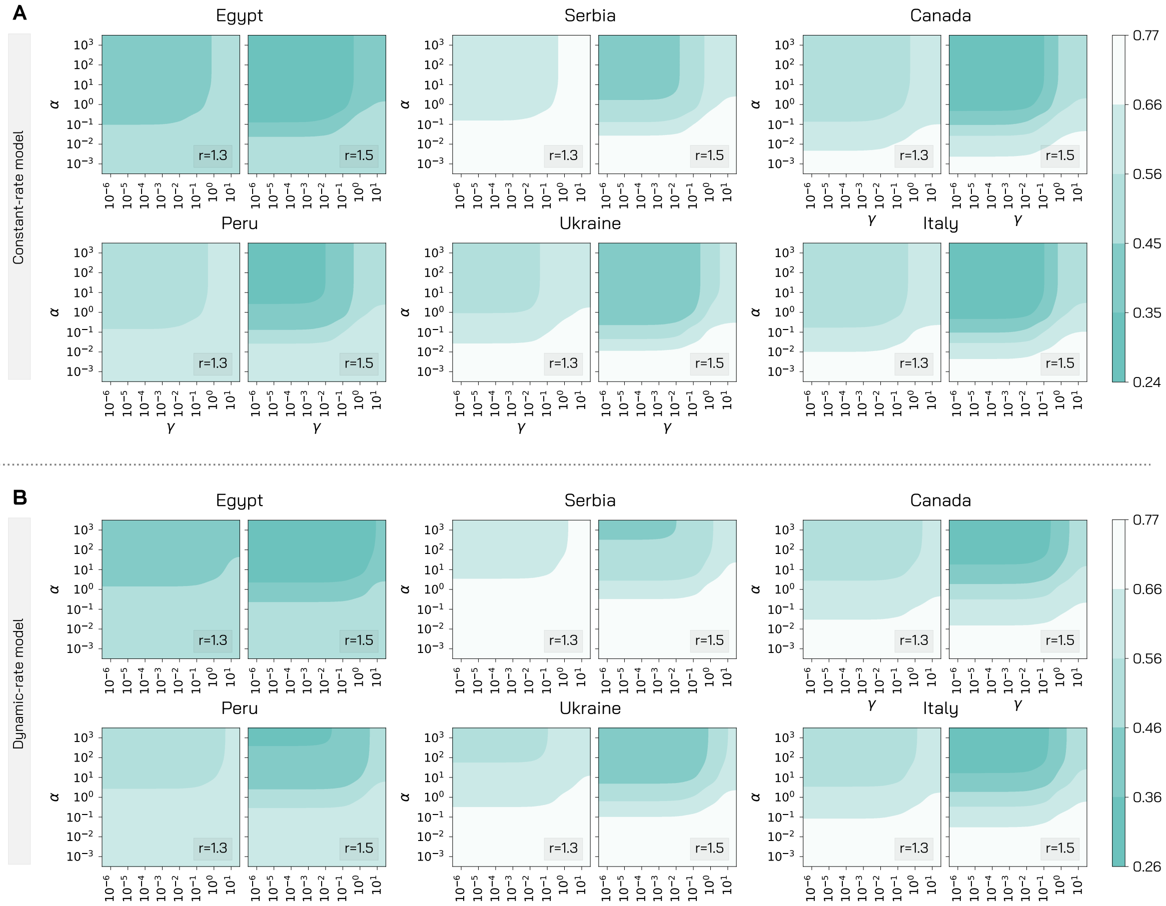


Figure 1: **Impact of model parameters governing the behavioural response.** For the six countries, we explore different values of $\alpha$ and $\gamma$ in terms of fraction of averted deaths with respect to a baseline without vaccine and no behavioural response (i.e. $r_V = 0\%$, $\alpha, \gamma = 0$). We consider two different values of the parameter $r$ (1.3 and 1.5) and we set $r_V = 1\%$ and $VE = 90\%$ and we employ vaccine strategy 1. In panel A we consider the constant rate mechanism to regulate behavioural transitions, while in panel B we use the dynamic-rate one (presented in the main text).

#### 2 Symptomatic cases

In the main text, we considered the number of deaths as a primary endpoint to evaluate the efficacy of behaviour and vaccine. In Figure 2 we compare the different vaccination strategies both in terms of averted deaths and averted symptomatic cases. We set $\alpha = 0$, therefore we do not consider behaviour change. We observe that the strategy prioritizing the elderly (i.e., strategy 1), is actually the most efficient one in reducing the number of deaths across the different population pyramids and contact patterns considered. The strategy prioritizing age groups $20 - 49$ (i.e., strategy 3) is the best one in reducing symptomatic cases for Serbia, Ukraine, Canada, and Italy. In the case of Egypt and Peru, while strategy 3 is preferable to strategy 1 when considering the fraction of averted symptomatic cases, the most efficient one in this case is the strategy that targets homogeneously the population (i.e., strategy 2). This may be due to the high contacts activity of individuals aged under 20, who are partly vaccinated since the beginning of the campaign when strategy 2 is employed.

In Figure 3, we repeat the analysis presented in the main text considering the relative symptomatic cases difference instead of the relative deaths difference as an endpoint to evaluate the effects of vaccines and behaviour on the spreading. With relative symptomatic cases difference, we simply intend the fraction of averted symptomatic cases in the presence of a vaccine with respect to baseline without vaccine. We consider the three vaccine prioritization strategies, the three vaccine efficacy, $VE = 50\%, 70\%, 90\%$, and different intensity of the behavioural responses by exploring a range of $\alpha$ values. We observe that the most efficient strategies at reducing the number of symptomatic cases are strategy 3 (for Serbia, Ukraine, Canada, and Italy) and 2 (for Egypt and Peru). Across the different countries, we observe that strategy 1 is generally the worst one in terms of averted symptomatic cases and it is also more affected by stronger behavioural responses.

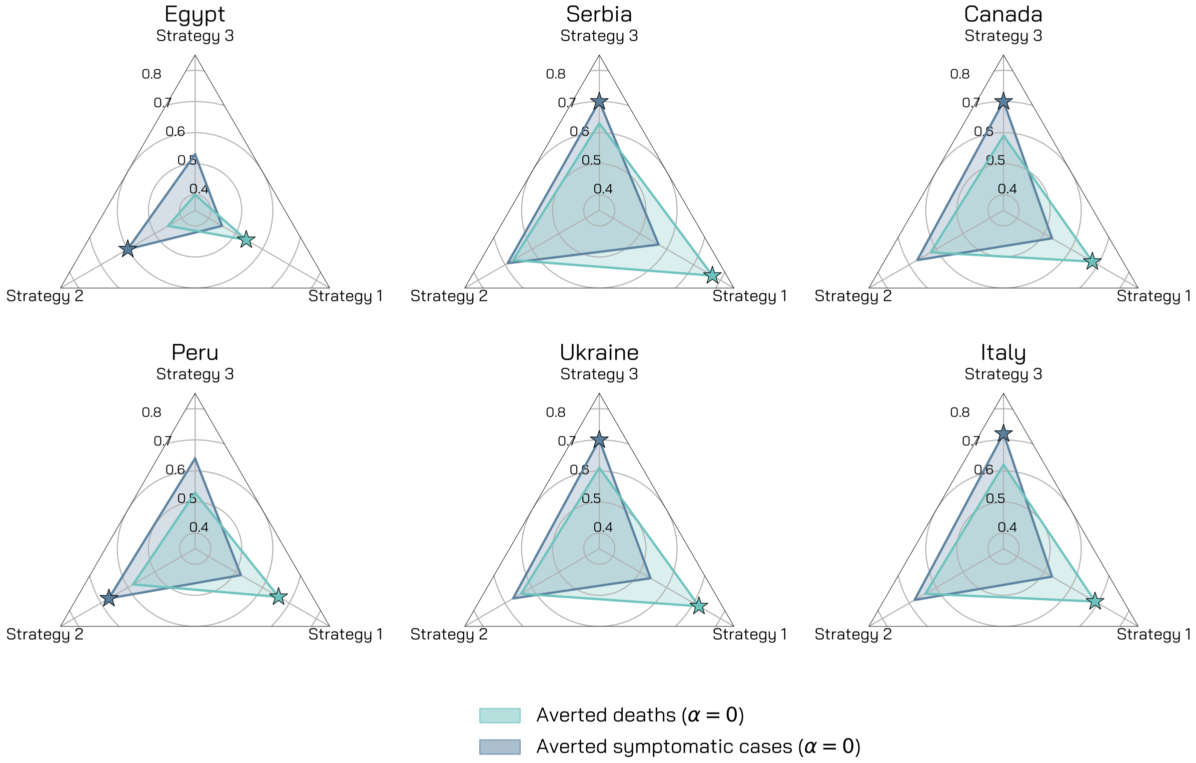


Figure 2: **Comparison of different vaccination strategies.** We compare the three vaccination strategies considered in terms of averted deaths and averted symptomatic cases when $\alpha = 0$ with respect to a baseline without vaccine. We set $\gamma = 0.5$, $R_0 = 1.15$, $r = 1.5$, $r_V = 1\%$, $VE = 90\%$, 1% of initially infected, 10% of initially immune individuals, and simulations length is set to 1 year.

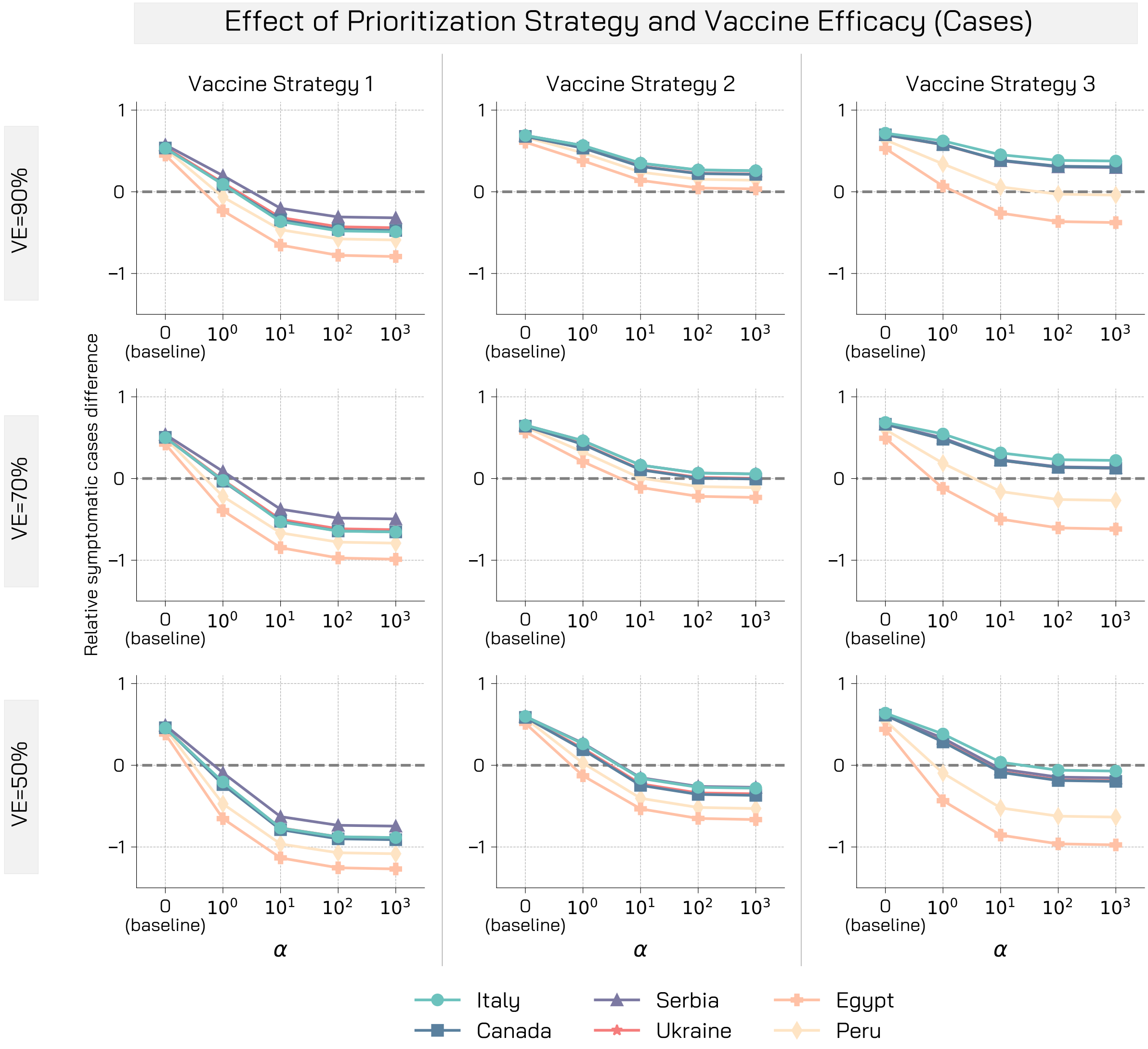


Figure 3: **Relative symptomatic cases difference for different vaccine efficacies and prioritization strategies.** Relative symptomatic cases difference is computed as the fraction of symptomatic cases that are avoided with a vaccine with respect to a baseline simulation without vaccine (and thus no behavioural response). We display results of the simulations for three vaccine efficacy and prioritization strategies. Other parameters used are $\gamma = 0.5$, $R_0 = 1.15$, $r = 1.5$, $r_V = 1\%$, 1% of initially infected, 10% of initially immune individuals, and simulations length is set to 1 year.

### 3 Calibration - Posterior Distributions

In Figure 4 we represent for the different countries the posterior distributions of the parameters calibrated through the Approximate Bayesian Computation rejection algorithm. In particular, we display the posterior distribution for the transmission parameter $\beta$, the initial number of infected individuals per $100,000$ individuals (split between the $L$, $P$, $I$, and $A$ compartments), and the delay in deaths $\Delta$ (i.e., the number of days between the transitions $R_I \to D$ and $D \to D^o$).

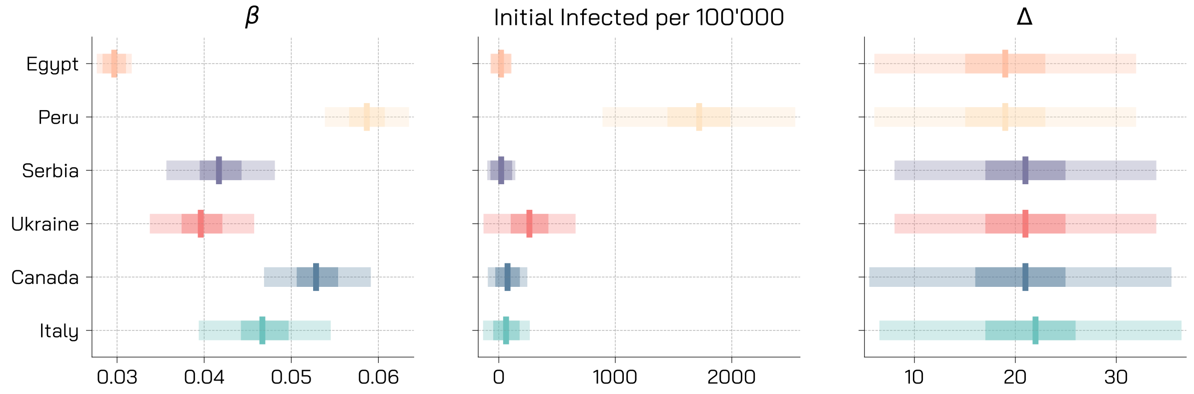


Figure 4: **Posterior Distributions.** We represent with boxplots the posterior distribution of the calibrated parameters for the different countries. Solid vertical lines indicate the median. The bounds of the darker shaded area indicate the first and third quartile, $Q1$, $Q3$. Their difference is defined as interquartile range $IQR = Q3 - Q1$. The bounds of the lighter shaded area indicate the "minimum" and the "maximum", defined respectively as $Q1 - 1.5IQR$ and $Q3 + 1.5IQR$.

### 4 Real Vaccination Rollout

We extend here the results presented in the main text for the calibrated model integrating also real data from an ongoing vaccination campaign. Because of data availability at the moment of writing, for this analysis, we will focus only on the case of Italy. We consider the daily number of first doses administered to individuals in the different age brackets from Ref. [1]. Indeed, all COVID-19 vaccines currently available in Italy require two injections. For the sake of simplicity and to match the available data with the modeling framework presented in this work, we consider only first doses administered daily. In doing so, we have a data-driven, time-varying estimate of the vaccine rollout speed $r_V$ and of the prioritization strategy. At the bottom of Figure 5 we represent the evolution in the first three months of 2021 of the cumulative percentage of people who received at least one dose. During this period, Italy managed to deliver at least one dose to roughly 10% of the population. In the figure, we also show the distribution of doses among the different age groups. Around 34% of the population aged over 75 received at least one dose, but we observe non-negligible percentages also in younger groups (for example more than 10% in the population aged $50-59$).

We consider the same calibration step presented in the main text which takes into account real epidemiological and mobility from Ref. [2, 3] during the period 2020/09/01 − 2020/12/31 to set the values of the free parameters of the model. After the calibration, we simulate the unfolding of the epidemic, of the restrictions, and of the vaccination campaign using the real rollout data between 2021/01/01 and 2021/03/22 (week 11). At the top of Figure 5, we represent the relative deaths difference for a spectrum of $\alpha$ values and for two values of the parameter $r$ capturing the increased risk of non-compliant individuals. Also when considering real data on the rollout progression, our findings are qualitatively similar to those obtained in the main text. Indeed, we observe that, while with an $\alpha = 0$ the fraction of averted deaths is about 11%, this fraction lowers and turns negative for higher values of $\alpha$. As expected, the higher value of $r$ leads to worse outcomes.

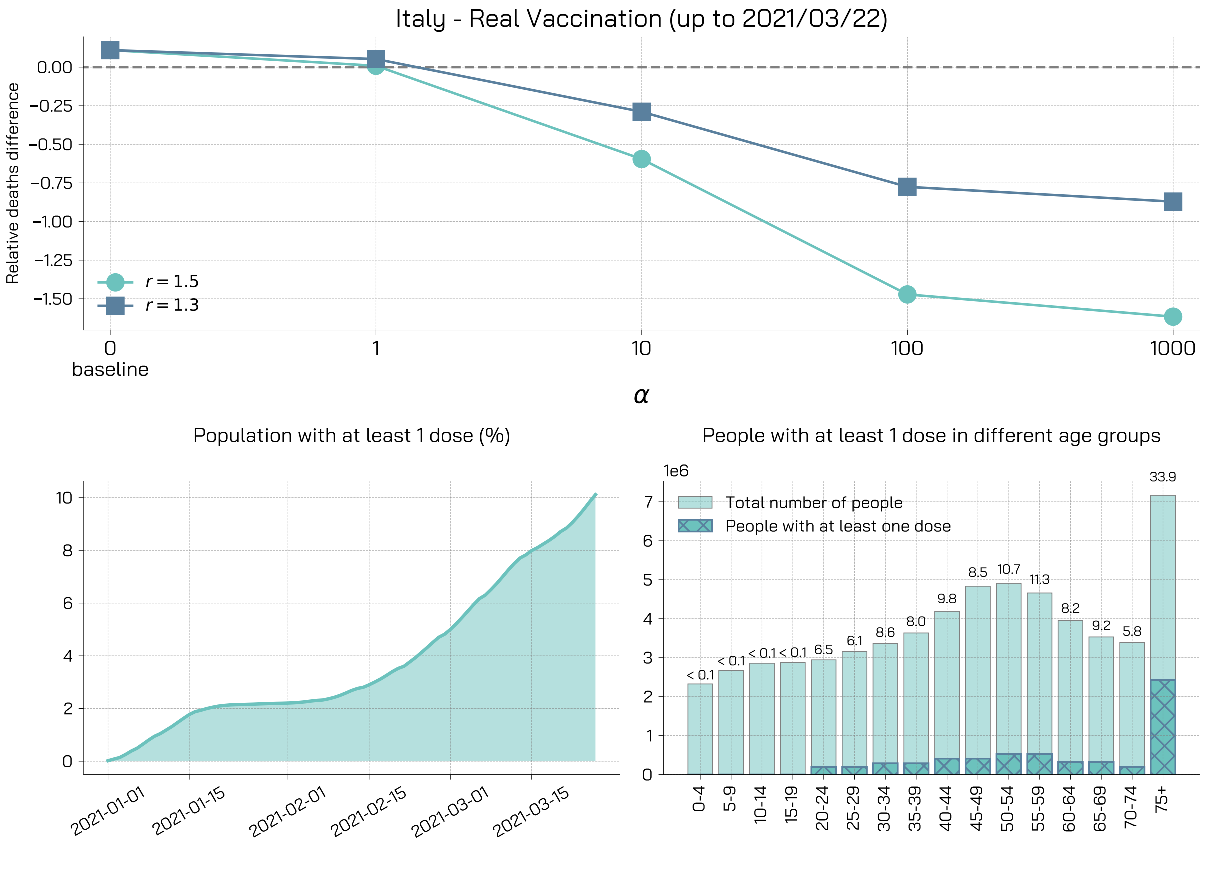


Figure 5: **Calibrated model with real vaccinations data.**

### 5 Reopening Scenarios

In the main text, we used the Google Mobility Report [2] and the Oxford Coronavirus Government Response Tracker [3] to inform contacts reduction over time up to week 11, 2021. Afterwards, we kept contacts at the level observed for week 11. Here, we propose a different reopening scenario in which we partially relax some of the measures. In particular, contacts at *work* and in *other locations* are increased by 25% with respect to the mixing levels observed in week 11, 2021. Additionally, we also consider a relaxation of the measures in schools subtracting 1 to the Oxford Coronavirus Government Response school index observed in week 11, 2021 (we check that schools are not already fully opened, in that case we do not change contacts in this setting).

We repeat the analysis presented in the main text with this new contacts scenario and we display results in Figure 6. In panel A, we observe that, as expected, after week 11 contacts increase in all countries, reaching in the case of Egypt almost the baseline level (with no restrictions). In panel B, we report the relative deaths difference for the three vaccination strategies and the two rollout speeds. We observe similar trends to those obtained in the main text. Indeed, a stronger behavioural response causes an increase in deaths otherwise averted thanks to the vaccine, and the faster rollout is much more robust than the slower one. Nonetheless, the overall effect of behaviour on averted deaths seems to be smaller. In the main text, in the worst possible case the relative deaths difference was about $-1.6$, while here is $-0.9$. This may seems counter-intuitive at first, since we are considering a more permissive contacts scenario. However, we underline that also the baseline simulation is changed respect to the analysis presented in the main text. Therefore, a smaller relative increase in deaths due to behaviour relaxation in the more permissive scenario can imply a higher absolute increase in deaths with respect to the more conservative one.

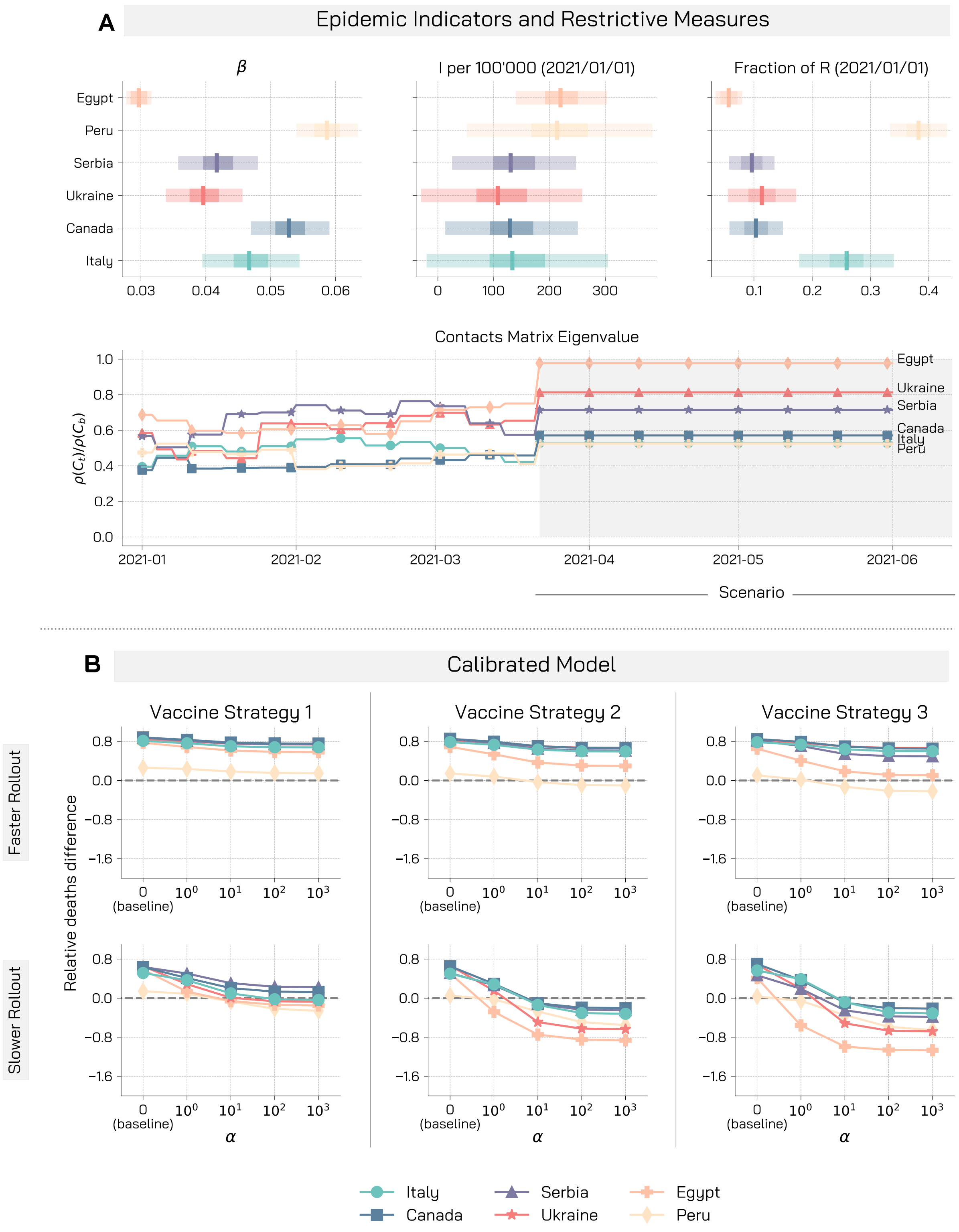


Figure 6: **Giving up NPIs during rollout may nullify the benefits brought by the vaccine - More permissive scenario.** A) We display for the different countries the boxplot of the calibrated infection parameter $\beta$, the projected number of symptomatic infectious cases per $100'000$ and the fraction of recovered at the 2021/01/01, start of the vaccination campaign in our simulation. We also report the ratio between the leading eigenvalue of the contacts matrix considering restrictions and of the baseline contacts matrix with no restrictions. B) We display the median relative deaths difference for the calibrated model in the different countries. We consider the three vaccination strategies and two possible rollout speed: $r_V = 1\%$ (faster rollout), and $r_V = 0.25\%$ (slower rollout). We run the model over the period 2021/01/01-2021/06/01. Other parameters are $\gamma = 0.5$, $r = 1.5$, $VE = 90\%$.

#### 6 Sensitivity Analysis: Behavioural Parameter $\gamma$

In the main text we generally kept constant the behavioural parameter $\gamma$ ($\gamma = 0.5$) and we let vary the other parameter $\alpha$. We informed the choice of $\gamma$ looking at the maximum number of COVID-19 deaths observed on a single day in the different countries. In the case of Italy, for example, $\gamma = 0.5$ would have implied a 60% probability for non-compliant to go back to safer behaviours with 1000 deaths in the previous step. Here, we report results obtained for different values of $\gamma$. We consider two additional values, $\gamma = 0.05$ and $\gamma = 2$, which in the previous example would imply respectively a return probability of 10% and 95%. In Figure 7 we compare, for each country, the relative deaths difference for the three values of $\gamma$ (0.05, 0.5, 2). We observe that, while the decreasing trend is common across the different $\gamma$ considered, curves for higher values of $\gamma$ are shifted upwards. In other words, when $\gamma$ grows the fraction of averted deaths increases. This is expected: indeed, a higher $\gamma$ implies greater awareness to deaths increase, and thus individuals go back to safer behaviours faster.

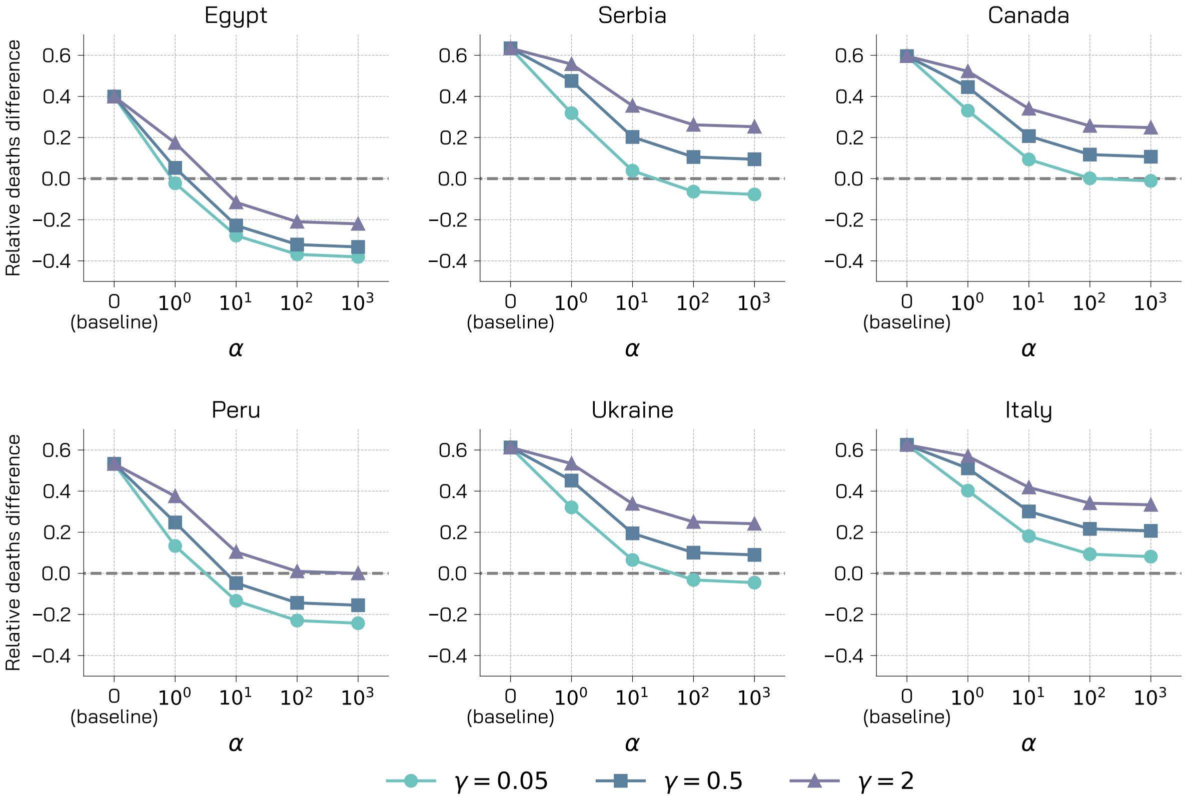


Figure 7: **Relative deaths difference for different $\gamma$.** We represent for the different countries the relative deaths difference for three values of the behavioural parameter $\gamma$ ($\gamma = 0.05, 0.5, 2$). We consider vaccination strategies prioritizing age groups $20 - 49$. We set $R_0 = 1.15$, $r = 1.5$, $r_V = 1\%$, $VE = 90\%$, 1% of initially infected, 10% of initially immune individuals, and simulations length is set to 1 year.

#### 7 $R_0$ calculation

We compute the basic reproduction number $R_0$ of the model proposed using the Next Generation Matrix method [4]. We consider the $4K$ equations that describe the evolution in time of the number of infected individuals $L$, $P$, $A$, and $I$:

$$\begin{aligned}
\frac{dL_k}{dt} =& + \beta \sum_{k'=1}^{K} C_{kk'} \frac{I_{k'} + \chi(P_{k'} + A_{k'})}{N_{k'}} S_k + r\beta \sum_{k'=1}^{K} C_{kk'} \frac{I_{k'} + \chi(P_{k'} + A_{k'})}{N_{k'}} S_k^{NC} + \\
&+ \beta(1-VE) \sum_{k'=1}^{K} C_{kk'} \frac{I_{k'} + \chi(P_{k'} + A_{k'})}{N_{k'}} V_k + r\beta(1-VE) \sum_{k'=1}^{K} C_{kk'} \frac{I_{k'} + \chi(P_{k'} + A_{k'})}{N_{k'}} V_k^{NC} - \epsilon L_k \\
\frac{dP_k}{dt} =& \epsilon L_k - \omega f P_k - \omega(1-f) P_k \\
\frac{dA_k}{dt} =& \omega f P_k - \mu A_k \\
\frac{dI_k}{dt} =& \omega(1-f) P_k - \mu(1 - IFR_k) I_k - \mu IFR_k I_k
\end{aligned} \tag{1}$$

In matrix notation:

$$\begin{bmatrix} \frac{dL_1}{dt} \\ \vdots \\ \frac{dL_K}{dt} \\ \frac{dP_1}{dt} \\ \vdots \\ \frac{dP_K}{dt} \\ \frac{dA_1}{dt} \\ \vdots \\ \frac{dA_K}{dt} \\ \frac{dI_1}{dt} \\ \vdots \\ \frac{dI_K}{dt} \end{bmatrix} = \begin{bmatrix} \beta \sum_{k'=1}^{K} C_{1k'} \frac{I_{k'}+\chi(P_{k'}+A_{k'})}{N_{k'}} S_1 + \cdots + r\beta(1-VE) \sum_{k'=1}^{K} C_{1k'} \frac{I_{k'}+\chi(P_{k'}+A_{k'})}{N_{k'}} V_1^{NC} \\ \vdots \\ \beta \sum_{k'=1}^{K} C_{Kk'} \frac{I_{k'}+\chi(P_{k'}+A_{k'})}{N_{k'}} S_K + \cdots + r\beta(1-VE) \sum_{k'=1}^{K} C_{Kk'} \frac{I_{k'}+\chi(P_{k'}+A_{k'})}{N_{k'}} V_K^{NC} \\ 0 \\ \vdots \\ 0 \\ 0 \\ \vdots \\ 0 \\ 0 \\ \vdots \\ 0 \end{bmatrix} + \tag{2}$$

$$- \begin{bmatrix} \epsilon L_1 \\ \vdots \\ \epsilon L_K \\ \omega P_1 - \epsilon L_1 \\ \vdots \\ \omega P_K - \epsilon L_K \\ \mu A_1 - \omega f P_1 \\ \vdots \\ \mu A_K - \omega f P_K \\ \mu I_1 - \omega(1-f) P_1 \\ \vdots \\ \mu I_K - \omega(1-f) P_K \end{bmatrix}$$

$$
\begin{bmatrix} \frac{d\theta_1}{dt} \\ \vdots \\ \frac{d\theta_K}{dt} \\ \frac{d\theta_{K+1}}{dt} \\ \vdots \\ \frac{d\theta_{2K}}{dt} \\ \frac{d\theta_{2K+1}}{dt} \\ \vdots \\ \frac{d\theta_{3K}}{dt} \\ \frac{d\theta_{3K+1}}{dt} \\ \vdots \\ \frac{d\theta_{4K}}{dt} \end{bmatrix} = \begin{bmatrix} F_1 \\ \vdots \\ F_K \\ 0 \\ \vdots \\ 0 \\ 0 \\ \vdots \\ 0 \\ 0 \\ \vdots \\ 0 \end{bmatrix} - \begin{bmatrix} V_1 \\ \vdots \\ V_K \\ V_{K+1} \\ \vdots \\ V_{2K} \\ V_{2K+1} \\ \vdots \\ V_{3K} \\ V_{3K+1} \\ \vdots \\ V_{4K} \end{bmatrix} \tag{3}
$$

Then, we define the disease free equilibrium (DFE) for age group $k$ as:

$$
(S_k, S_k^{NC}, V_k, V_k^{NC}, L_k, P_k, A_k, I_k, R_k, D_k, D_k^o) = (N_k, 0, 0, 0, 0, 0, 0, 0, 0, 0, 0) \tag{4}
$$

Indeed, the vaccination (and thus the behavioural dynamics) starts only after time $t_V$ and when $t_V = 0$ the number of vaccinated in the early stage of the dynamics is small (here, at most 1% of the population is vaccinated daily). We also assume that $R_k << N_k$. We define the two matrices $F$ and $V$ as follows: $F_{ij} = \frac{dF_i}{d\theta_j}\Big|_{DFE}$ and $V_{ij} = \frac{dV_i}{d\theta_j}\Big|_{DFE}$. These can be written as:

$$
F = \begin{bmatrix}
0 & \cdots & 0 & \beta\frac{N_1C_{11}\chi}{N_1} & \cdots & \beta\frac{N_1C_{1K}\chi}{N_K} & \beta\frac{N_1C_{11}\chi}{N_1} & \cdots & \beta\frac{N_1C_{1K}\chi}{N_K} & \beta\frac{N_1C_{11}}{N_1} & \cdots & \beta\frac{N_1C_{1K}}{N_K} \\
\vdots & \ddots & \vdots & \vdots & \ddots & \vdots & \vdots & \ddots & \vdots & \vdots & \ddots & \vdots \\
0 & \cdots & 0 & \beta\frac{N_KC_{K1}\chi}{N_1} & \cdots & \beta\frac{N_KC_{KK}\chi}{N_K} & \beta\frac{N_KC_{K1}\chi}{N_1} & \cdots & \beta\frac{N_KC_{KK}\chi}{N_K} & \beta\frac{N_KC_{K1}}{N_1} & \cdots & \beta\frac{N_KC_{KK}}{N_K} \\
0 & \cdots & 0 & 0 & \cdots & 0 & 0 & \cdots & 0 & 0 & \cdots & 0 \\
\vdots & \ddots & \vdots & \vdots & \ddots & \vdots & \vdots & \ddots & \vdots & \vdots & \ddots & \vdots \\
0 & \cdots & 0 & 0 & \cdots & 0 & 0 & \cdots & 0 & 0 & \cdots & 0
\end{bmatrix} \tag{5}
$$

$$
V = \begin{bmatrix}
\epsilon & \cdots & 0 & 0 & \cdots & 0 & 0 & \cdots & 0 & 0 & \cdots & 0 \\
\vdots & \ddots & \vdots & \vdots & \ddots & \vdots & \vdots & \ddots & \vdots & \vdots & \ddots & \vdots \\
0 & \cdots & \epsilon & 0 & \cdots & 0 & 0 & \cdots & 0 & 0 & \cdots & 0 \\
-\epsilon & \cdots & 0 & \omega & \cdots & 0 & 0 & \cdots & 0 & 0 & \cdots & 0 \\
\vdots & \ddots & \vdots & \vdots & \ddots & \vdots & \vdots & \ddots & \vdots & \vdots & \ddots & \vdots \\
0 & \cdots & -\epsilon & 0 & \cdots & \omega & 0 & \cdots & 0 & 0 & \cdots & 0 \\
0 & \cdots & 0 & -\omega f & \cdots & 0 & \mu & \cdots & 0 & 0 & \cdots & 0 \\
\vdots & \ddots & \vdots & \vdots & \ddots & \vdots & \vdots & \ddots & \vdots & \vdots & \ddots & \vdots \\
0 & \cdots & 0 & 0 & \cdots & -\omega f & 0 & \cdots & \mu & 0 & \cdots & 0 \\
0 & \cdots & 0 & -\omega(1-f) & \cdots & 0 & 0 & \cdots & 0 & \mu & \cdots & 0 \\
\vdots & \ddots & \vdots & \vdots & \ddots & \vdots & \vdots & \ddots & \vdots & \vdots & \ddots & \vdots \\
0 & \cdots & 0 & 0 & \cdots & -\omega(1-f) & 0 & \cdots & 0 & 0 & \cdots & \mu
\end{bmatrix} \tag{6}
$$

The basic reproduction number is defined as $R_0 = \rho(FV^{-1})$, where $\rho(\cdot)$ indicates the spectral radius. First, we compute $V^{-1}$. We recognize that $V$ and $F$ can be written in blocks as:

$$
F = \begin{bmatrix} 0 & \chi\beta\tilde{C} & \chi\beta\tilde{C} & \beta\tilde{C} \\ 0 & 0 & 0 & 0 \\ 0 & 0 & 0 & 0 \\ 0 & 0 & 0 & 0 \end{bmatrix}, V = \begin{bmatrix} H & 0 & 0 & 0 \\ I & L & 0 & 0 \\ 0 & M & N & 0 \\ 0 & P & 0 & R \end{bmatrix} \tag{7}
$$

Where all the block components of $V$ are diagonal matrices, and $\tilde{C}$ is the contacts matrix weighted by the relative population in different age groups (i.e. $\tilde{C_{ij}} = \frac{N_i}{N_j}C_{ij}$). The inverse of a block matrix $\begin{bmatrix} A & 0 \\ C & D \end{bmatrix}$

can be written as $\begin{bmatrix} A^{-1} & 0 \\ -D^{-1}CA^{-1} & D^{-1} \end{bmatrix}$, where in our case $A = \begin{bmatrix} H & 0 \\ I & L \end{bmatrix}$, $C = \begin{bmatrix} 0 & M \\ 0 & P \end{bmatrix}$, $D = \begin{bmatrix} N & 0 \\ 0 & R \end{bmatrix}$. Therefore, we compute:

$$A^{-1} = \begin{bmatrix} H & 0 \\ I & L \end{bmatrix}^{-1} = \begin{bmatrix} H^{-1} & 0 \\ -L^{-1}IH^{-1} & L^{-1} \end{bmatrix} \tag{8}$$

$$D^{-1} = \begin{bmatrix} N & 0 \\ 0 & R \end{bmatrix}^{-1} = \begin{bmatrix} N^{-1} & 0 \\ 0 & R^{-1} \end{bmatrix} \tag{9}$$

$$\begin{aligned} -D^{-1}CA^{-1} &= -\begin{bmatrix} N^{-1} & 0 \\ 0 & R^{-1} \end{bmatrix} \begin{bmatrix} 0 & M \\ 0 & P \end{bmatrix} \begin{bmatrix} H^{-1} & 0 \\ -L^{-1}IH^{-1} & L^{-1} \end{bmatrix} \\ &= -\begin{bmatrix} 0 & N^{-1}M \\ 0 & R^{-1}P \end{bmatrix} \begin{bmatrix} H^{-1} & 0 \\ -L^{-1}IH^{-1} & L^{-1} \end{bmatrix} \\ &= \begin{bmatrix} N^{-1}ML^{-1}IH^{-1} & -N^{-1}ML^{-1} \\ R^{-1}PL^{-1}IH^{-1} & -R^{-1}PL^{-1} \end{bmatrix} \end{aligned} \tag{10}$$

Substituting these expressions, we can then write $V^{-1}$:

$$V^{-1} = \begin{bmatrix} H^{-1} & 0 & 0 & 0 \\ L^{-1}IH^{-1} & L^{-1} & 0 & 0 \\ N^{-1}ML^{-1}IH^{-1} & -N^{-1}ML^{-1} & N^{-1} & 0 \\ R^{-1}PL^{-1}IH^{-1} & -R^{-1}PL^{-1} & 0 & R^{-1} \end{bmatrix} \tag{11}$$

The next step consists in computing the product $FV^{-1}$:

$$FV^{-1} = \begin{bmatrix} 0 & \chi\beta\tilde{C} & \chi\beta\tilde{C} & \beta\tilde{C} \\ 0 & 0 & 0 & 0 \\ 0 & 0 & 0 & 0 \\ 0 & 0 & 0 & 0 \end{bmatrix} \begin{bmatrix} H^{-1} & 0 & 0 & 0 \\ L^{-1}IH^{-1} & L^{-1} & 0 & 0 \\ N^{-1}ML^{-1}IH^{-1} & -N^{-1}ML^{-1} & N^{-1} & 0 \\ R^{-1}PL^{-1}IH^{-1} & -R^{-1}PL^{-1} & 0 & R^{-1} \end{bmatrix}$$

$$= \beta\tilde{C} \begin{bmatrix} (-\chi\mathbb{1} + \chi N^{-1}M + R^{-1}P)L^{-1}IH^{-1} & \chi L^{-1} - \chi N^{-1}ML^{-1} - R^{-1}PL^{-1} & \chi N^{-1} & R^{-1} \\ 0 & 0 & 0 & 0 \\ 0 & 0 & 0 & 0 \\ 0 & 0 & 0 & 0 \end{bmatrix} \tag{12}$$

Finally, we are left with finding the spectral radius of $FV^{-1}$ (i.e., its largest eigenvalue). The eigenvalue problem can be written as $det(FV^{-1} - \lambda\mathbb{1}) = 0$. Given the structure of $FV^{-1}$, and since we are interested in non-trivial solutions ($\lambda \neq 0$), the problem reduces to:

$$det[\beta\tilde{C}(-\chi\mathbb{1} + \chi N^{-1}M + R^{-1}P)L^{-1}IH^{-1} - \lambda\mathbb{1}] = 0 \tag{13}$$

Since $N$, $M$, $R$, $P$, $L$, $I$, and $H$ are all diagonal we easily compute the inverses and products and simplify the expression to:

$$det\left[\beta\left(\frac{\chi}{\omega} + \frac{f\chi}{\mu} + \frac{1-f}{\mu}\right)\tilde{C} - \lambda\mathbb{1}\right] = 0 \tag{14}$$

Therefore, finding the spectral radius of $FV^{-1}$ is equivalent to solving the eigenvalue problem for $\beta\left(\frac{\chi}{\omega} + \frac{f\chi}{\mu} + \frac{1-f}{\mu}\right)\tilde{C}$ and taking the largest eigenvalue. Finally, we have that $R_0 = \beta\left(\frac{\chi}{\omega} + \frac{f\chi}{\mu} + \frac{1-f}{\mu}\right)\rho(\tilde{C})$

[4] Pauline Driessche. Reproduction numbers of infectious disease models. *Infectious Disease Modelling*, 2, 06 2017.
